# Low levels of Fe and Se with high IL-6/IL-10 likely influence nutritional immunity in tuberculosis patients

**DOI:** 10.1101/2021.09.09.21263312

**Authors:** Sandeep R. Kaushik, Sukanya Sahu, Hritusree Guha, Sourav Saha, Ranjit Das, Rukuwe-u Kupa, Wetetsho Kapfo, Trinayan Deka, Rumi Basumatary, Asunu Thong, Arunabha Dasgupta, Bidhan Goswami, Amit Kumar Pandey, Lahari Saikia, Vinotsole Khamo, Anjan Das, Ranjan Kumar Nanda

## Abstract

Tuberculosis (TB) patients present dysregulated immunity, iron metabolism and anaemia of inflammation. In this study, circulatory cytokines, trace metals, and iron-related proteins (hepcidin, ferroportin, transferrin, DMT1, Nramp1, ferritin, ceruloplasmin, hemojuvelin, aconitase, transferring receptor) were monitored in case (active tuberculosis patients: ATB) and control (non-tuberculosis: NTB and healthy) study populations (n=72, male, 42.94 mean age (16-83)). Using serum elemental and cytokine levels, a partial least square discriminate analysis model (PLS-DA) was built and variables with a VIP score of >0.6 were selected as important markers. A biosignature of IL-13, IL-12(p70), IFN-γ, IL-10, IL-5, IL-18, IL-4, Selenium, and Aluminium clustered ATB away from controls. Interestingly, low iron and selenium levels, while high copper and aluminum levels were observed in ATB subjects. All the important serum cytokines were positively correlated in ATB subjects. A low abundance of transferrin, ferroportin, and hemojuvelin, while higher ferritin and ceruloplasmin levels explained an altered iron metabolism in ATB subjects which partially resolved upon completion of treatment. Further, the identified biosignature in TB patients, that explained anemia of inflammation, along with perturbed iron homeostasis could be useful targets for the development of host-directed adjunct therapeutics.

## Introduction

Tuberculosis (TB) is caused by *Mycobacterium tuberculosis (Mtb) infection* and is still a major killer worldwide.^1^ TB patients present with a dysregulated immune systems and release pro-inflammatory cytokines inducing a cascade of activities hampering iron homeostasis.^2^ Hepcidin from hepatocytes is released into the circulation which subsequently binds to ferroportin and induces its degradation through ubiquitylation.^3^ Increased ferroportin degradation leads to reduced circulatory iron and causes anemia of inflammation (AI).^2,4^ Further, during AI, even though host may have enough stored iron, it becomes unavailable for use and the dietary iron absorption is lowered as well. Moreover, as the metabolic crossroads of iron are also linked to copper, zinc, and selenium levels,^5,6^ and because of their heightened reactivity, intracellular levels of these trace metals are regulated critically by the pathogen. However, the host system exploits the indispensability and toxicity of these metals to safeguard itself from bacterial invaders.^7^ Nutritional immunity involves in the mechanism to withhold trace metals like iron to limit pathogen’s growth.^8^ Interlinking host immuno-profile with levels of circulatory trace metals and iron-related proteins, may help design newer approaches to develop adjunct therapeutics for TB. In this study, these important molecules were monitored in newly diagnosed TB patients, controls and followed up TB patients.

## Methodology

### Ethics statement and subject recruitment

This study was part of a project activity approved by the institutional review board and ethics committees of Agartala Government Medical College-Agartala (protocolF.4[6-9]/AGMC/Academic/IEC Committee/2015/8965, dated 25 April 2018), Nagaland Hospital Authority-Kohima (NHAK/HLRC/RES-3/2013/64), Assam Medical College-Dibrugarh (AMC/EC/PG/3530) and International Centre for Genetic engineering and Biotechnology (ICGEB) New Delhi (ICGEB/IEC/2014/07). After receiving signed informed consent, subjects presenting with 2 weeks of cough, fever, and weight loss at the outpatient departments of the partnering hospitals were recruited. Collected sputum samples were subjected to microscopy and, GeneXpert tests, and subjects with positive test results for both were grouped as active tuberculosis patients (ATB) while those with negative results were grouped as non-TB (NTB).^9^ NTB subjects were clinically diagnosed as asthma, COPD, lung cancer, and pneumonia patients. Only male subjects with negative HIV test results were selected. Healthy subjects without receiving any medication for at least one week before sampling were recruited as controls. Whole blood samples (4 mL) were collected in vacutainer tubes for serum isolation by centrifuging at 1,500 *g* for 10 mins at 4°C. Aliquots of serum were stored at -80°C till further analysis and a maximum of two freeze-thaw cycles were allowed.

### Serum cytokine, micronutrient and iron metabolizing portein profiling

Serum pro-inflammatory (interleukin (IL)-1β, IL-2, IL-6, IL-12, IFN-γ, TNF-α), anti-inflammatory cytokines (IL-4, IL-5, IL-10, and IL-13) and IL-18 levels were quantified using a Bioplex Microplate array reader (Bio-Rad Bio-Plex 200 systems, USA). Serum samples were digested using HNO_3_ and micronutrient levels were quantified using an Inductively Coupled Plasma Mass Spectrometry (ICP-MS). Iron metabolism-related proteins were probed in serum of study subjects using Western blot analysis. For detailed methodologies used for Bioplex, ICP-MS and Western blot analyses, refer to supplementary information.

### Statistical analysis

MetaboAnalyst 5.0 was used for building a Partial Least Square-Discriminate Analysis (PLS-DA) model and variables with a Variable Importance in Projection (VIP) score >0.6 were selected as important markers.^10^ (refer to supplementary information).

## Results and discussion

In this study, 72 age-matched male subjects from ATB (n=29, mean age 41.62), NTB (n=20, 46.30) and healthy (n=23, 41.43) groups were included (Fig. 1A, Table S1, Fig. S1). As circulatory iron levels significantly alter during the reproductive cycle of female subjects and thus excluded from the present study.^4,11^ A set of IL-13, IL-12(p70), IFN-γ, IL-10, IL-5, IL-18, IL-4, Se and Al qualified (as per criteria described above) as important markers from the PLS-DA analysis (Fig. 1B,1C). Circulatory Th_2_ cytokines (IL-4, IL-5, IL-10, IL-13) were strongly depressed in ATB subjects, corroborating earlier reports (Fig. 1D).^12^ Lower levels of IFN-γ and IL-10 were observed in ATB which have been reported to be directly linked to TB cure.^13^ Higher IL-18 and low IL-12 levels were observed in the ATB group (Fig. 1D). Reports showed higher Th_1_ cytokine levels in ATB subjects, but we observed lower levels of IFN-γ and IL-12p70 and rest (IL-1β, IL-2, IL-6, and TNF-α) remained similar in all study groups (Fig. S2). Interestingly, IL-6/IL10 and TNF-α/IL-10 levels were significantly higher compared to the healthy group (4.1 and 1.4 respectively p<0.05) (Fig. 1F). The serum IL-4 levels were positively correlated with IL-5, IL-10, IL-13, and IFN-γ levels in ATB and with IL-5 and, IL-18 in healthy groups (Fig. 1G). Significantly low serum selenium and high aluminum levels were observed in ATB subjects (Fig. 1E).^14^ Selenium participates in key enzymes involved in antioxidant activities, was found negatively correlated to aluminum in ATB subjects and to IL-10 in controls (Fig. 1G). Higher circulatory aluminum may saturate transferrin binding sites and impact haematopoiesis under hypoferric conditions.^15-16^ We found Aluminum levels to positively correlate with anti-inflammatory cytokines whereas negatively with pro-inflammatory cytokines (Fig. 1G). In ATB patients, we observed lower iron and higher copper levels but were statistically insignificant (Fig. S3) and an inverse relationship between serum IL-6 and iron levels(Fig. S4A). Rest of the elements were similar between all groups (Fig. S3). Through nutritional immunity, the host sequesters circulatory trace metals, including iron, to limit pathogenicity during infection and the observed changes in trace metal levels might be benefiting the host.^8-17^

**Figure 1:**
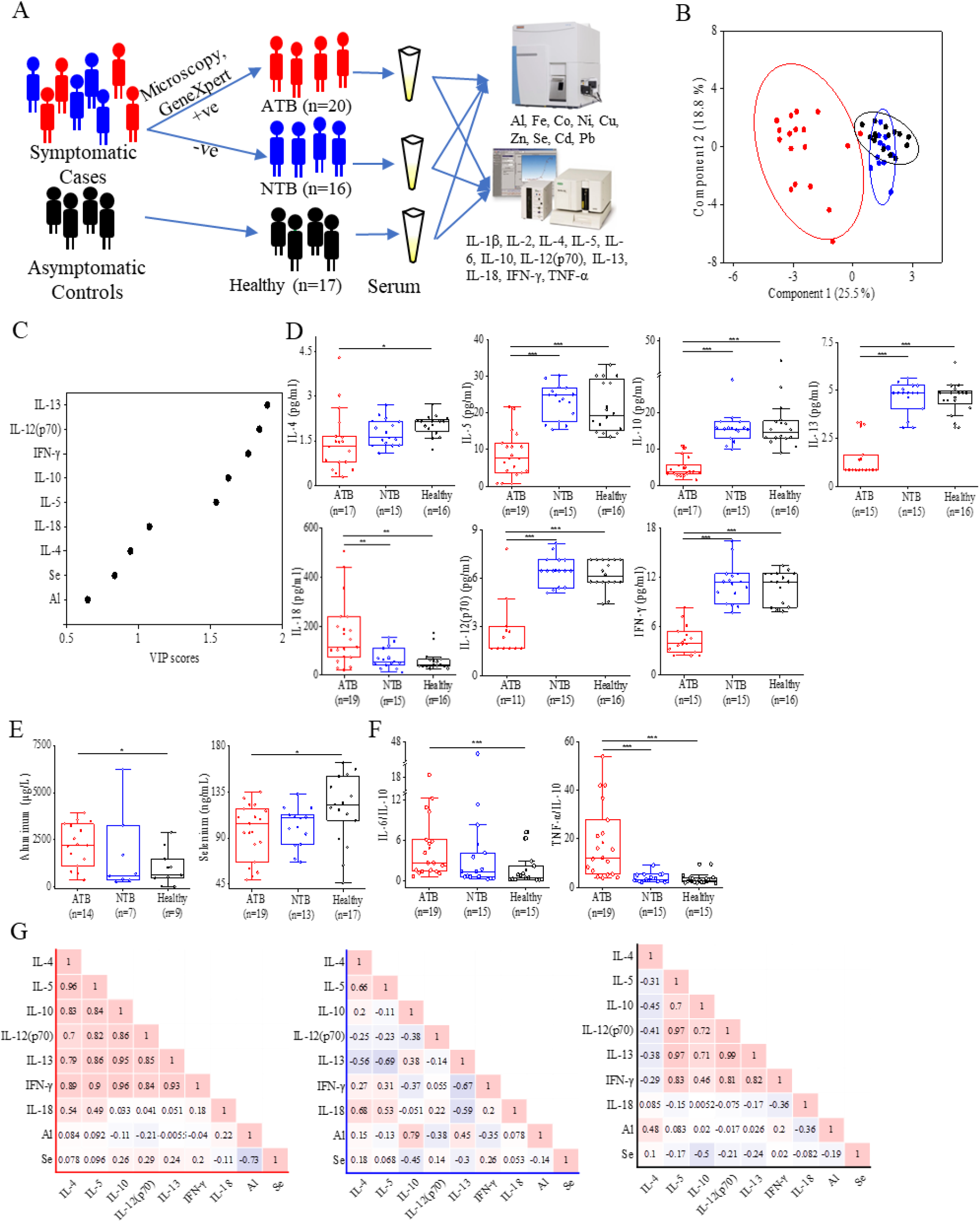
Serum cytokine and trace metal levels show significant differences between active tuberculosis (ATB), non-tuberculosis (NTB), and healthy controls. A) Outline of subject recruitment and classification for serum Bioplex and ICP-MS analyses; B) Partial least squares discriminant analysis (PLS-DA) of identified serum cytokines and trace metals showed ATB study group cluster away from NTB and healthy control subjects. C) Important cytokines and trace metals identified based on Variable Importance in Projection (VIP) score from the PLS-DA model. D) Individual serum cytokine levels, E) trace metal levels and F) IL-6/IL-10 and TNFα/IL-10 ratios in the study groups. G) Correlation (Pearson r) values between important markers in ATB, NTB, and Healthy study groups. (*: p<0.05, **: p<0.01, ***: p<0.001)

Both iron deficiency and iron overload are detrimental to the host and therefore, iron homeostasis is maintained within a narrow range. *Mtb* infection induces inflammation, lowers labile iron pool, and alters erythropoiesis requirements which may be contributed by differential expression of iron carrier, transporters, and storage proteins. IL-6 induces hepatic hepcidin production, but we observed low serum hepcidin levels in ATB patients (Fig. 2A,2B, Fig. S5).^18-22^ Interestingly, we observed significantly low levels of ferroportin in TB patients. Higher levels of ceruloplasmin (promotes Fe^2+^ oxidation in plasma) and ferritin (involved in iron storage) were observed in ATB subjects (Fig. 2A,2B, Fig. S6). Higher transferrin receptor and low transferrin levels in ATB subjects (Fig. 2A,2B, Fig. S5) may enhance holotransferrin-transferrin receptor complex formation to deplete the circulatory iron and transferrin. The levels of DMT1, an endosomal transmembrane transporter involved in cellular influx of iron and aconitase (IRP1), which may prevent intracellular pathogen’s growth, were found to be marginally lower in ATB subjects (Fig. 2A,2B, Fig. S5).^22-23^ Inflammation reduces hepatic hemojuvelin production and impacts iron sensing, and a similar observation was found in ATB patients.^24-25^ The levels of Nramp1 which transports phagosomal iron (Fe^2+^) to the cytosol was observed to be reduced in ATB subjects which may be facilitating *Mycobacterial* growth (Fig. 2A,2B, Fig. S5). A similar observation was found in study groups recruited from another site (Fig. S6,S7). A majority of these iron metabolizing proteins returned to the baseline in TB patients post-treatment (Fig. 2C,2D, Fig. S8). Compared to controls, a dysregulated iron metabolism was observed in ATB patients (Fig. 2E).

**Figure 2:**
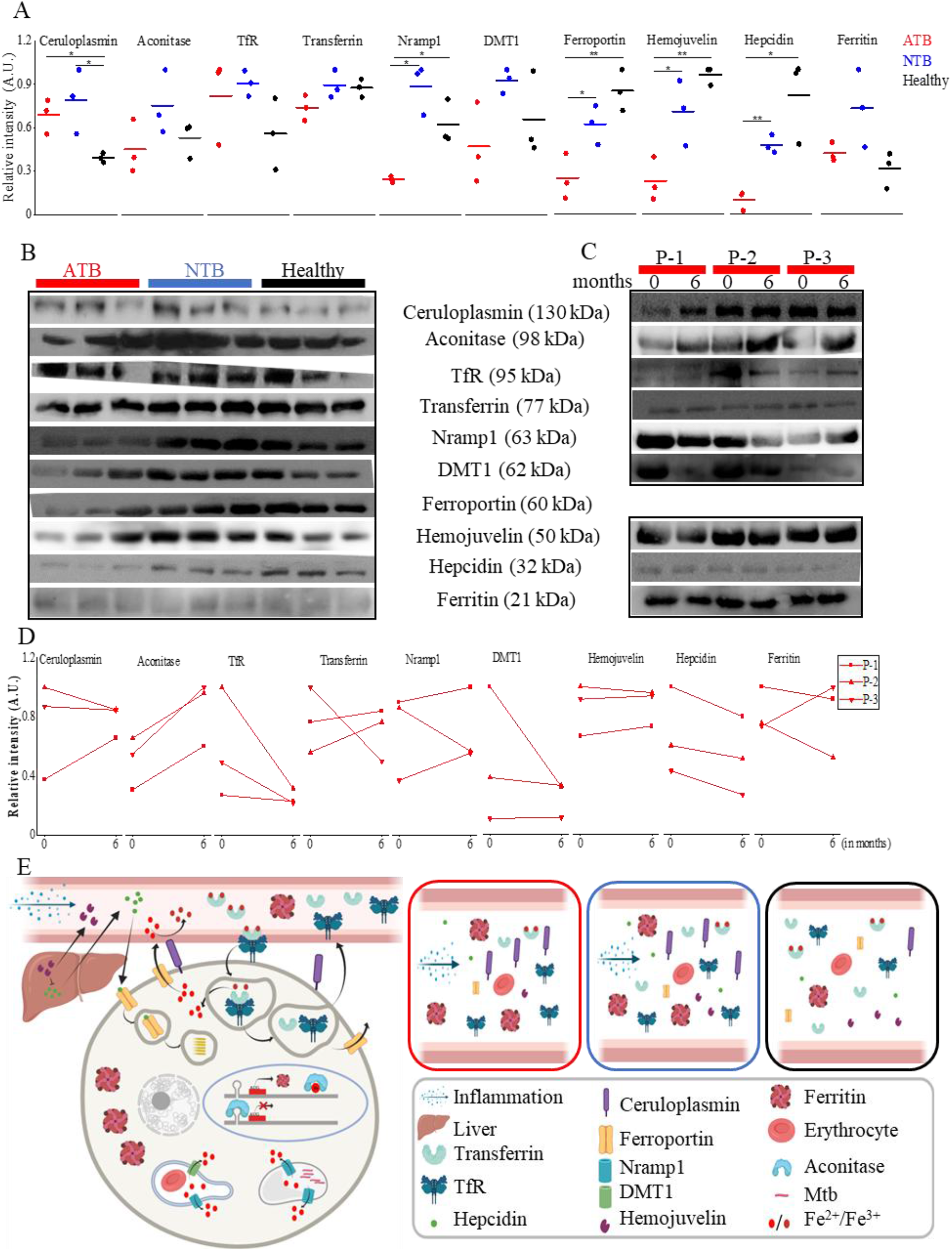
Expression levels of serum iron metabolism-related proteins in active tuberculosis (ATB), non-tuberculosis (NTB), and healthy control subjects. A) Iron metabolism-related proteins expression shows significant differences between ATB, NTB, and healthy controls. B) Western blot images used for calculating iron metabolism-related protein expression levels in ATB, NTB, and Healthy subjects. C) Western blot images of iron metabolizing proteins in longitudinally followed up tuberculosis patients at case presentation (0 months) and completion of treatment (6 months). D) Treatment-induced alteration in serum protein expression levels in longitudinally followed up tuberculosis patients. E) Dysregulated iron metabolism depicted in ATB/NTB/Healthy subjects. (P: patient; TfR: Transferrin receptor, DMT1: Divalent metal ion transporter 1; A.U.: arbitrary unit; *: p<0.05; **: p<0.01).

In conclusion, this study demonstrated a depressed Th_2_ profile and low serum selenium levels in TB patients. This report provides evidence for paradigms in nutrient metal homeostasis at the host-pathogen interface. The clinical benefits of selenium supplementation in TB patients need further validation. Additionally, the observed anemia of inflammation and dysregulated iron homeostasis in TB patients may provide a useful target for adjunct therapeutics development.

## Data Availability

All data referred in this article will be shared to the requesting readers with appropriate justification.

## Acknowledgments

We acknowledge the Department of Biotechnology (DBT), Government of India for supporting activities through research grants (BT/510/NE/TBP/2013 dated 11-08-2014 and MDR-TB/2017/39) and ICGEB New Delhi for providing access to ICP-MS facility and core support. Sandeep R. Kaushik received Senior Research Fellowship from the DBTand Sukanya Sahu received support from the Council of Scientific and Industrial Research (CSIR) New Delhi. All hospital staff involved in subject recruitment, testing, and classification are highly acknowledged. We thank Subia Akram for assisting in diagram preparation using BioRender.

## Authorship Contributions

SRK, SS carried out all the laboratory profiling experiments; study subject recruitment for classification was conducted by HG, RD, SS, RK, WK, TD, and RB under the guidance of AT, AD, BG, LS, VK, and AD; Funds for this work was generated by RKN, LS, VK and AD; AP shared resources and was involved in experimental planning; SRK and RKN wrote the first draft of the manuscript and revised it, incorporating the comments of all authors.

