## Supplementary information for "Low levels of Fe and Se with high IL-6/IL-10 likely influence nutritional immunity in tuberculosis patients"

**Supplemental Methodology:**

**Serum cytokine profiling:** Serum pro-inflammatory (interleukin (IL)-1β, IL-2, IL-6, IL-12, IFN-γ, TNF-α), anti-inflammatory cytokines (IL-4, IL-5, IL-10, and IL-13) and IL-18 levels were quantified using a Bioplex Microplate array reader (Bio-Rad Bio-Plex 200 systems, USA). Briefly, to the diluted serum samples, conjugated beads (Bio-Rad, USA) were added followed by biotinylated detection antibodies. After adding streptavidin to the test samples, standards, and blanks, the fluorescence intensity for all the bead regions was measured using a Bioplex Microplate array reader (Bio-Rad Bio-Plex 200 systems, USA).

**Micronutrient profiling in serum samples using Inductive Coupled Plasma Mass Spectrometry (ICP-MS):** Diluted serum samples (50 µl) in ultrapure water (Honeywell) were digested using $HNO_3$ (225711, Sigma, USA) and $H_2O_2$ (Supelco, 107298, Hydrogen peroxide 30% Suprapur®) in a Multiwave-Pro digestor (Anton Paar, USA) for 30 min at 140°C. Trace metal levels were quantified using an ICP-MS (Thermo Scientific iCAP-TQ) in Kinetic energy discrimination (KED) mode using Helium.[1,2]

**Western blot experiment:** Equal amount of serum proteins from study subjects were probed by Western blot analysis. Denatured serum proteins were loaded on SDS-PAGE gel for separation and transferred to PVDF membrane using Semi-dry transfer apparatus (TE77, semi dry apparatus, Amersham). The blots were incubated with primary antibody (Transferrin (15 µg); ab82411, Transferrin receptor (30 µg); ab1086, Ferroportin (20 µg); ab78066, Hepcidin (20 µg); ab30760, Dmt1 (30 µg); ab55735, Nramp1 (20 µg); ab59696, Aconitase (25 µg); ab126595, Hemojuvelin (15 µg); ab54431 from Abcam, USA and Ferritin (30 µg); D1D4 and Ceruloplasmin (15 µg); D7Q5W from Cell Signalling Technology USA) for overnight at 4°C using a shaker. After washing, the blots were incubated with secondary antibody (Anti-Rabbit, Sigma A-6154) for 2 hours and developed using Pico-Plus ECL (Pierce, Thermo Fisher) on X-ray films or imaging system (ChemiDoc MP Bio-Rad, USA). Image-J was used for densitometric calculations. Parallel gels were run using 5 µg serum protein and silver stained for every blot.

**Statistical analysis:** MetaboAnalyst 5.0 tool was used for Partial Least Square-Discriminate Analysis (PLS-DA) model building. Variables with a Variable Importance in Projection (VIP) score >0.6 were selected as important markers. OriginPro 2020b (64-bit) 9.7.5.184 (Student Version) was used for box plots, line plots, scatter plots and correlation analysis. Univariate statistical tools like Student's t-test (paired or unpaired) were performed to calculate the significance level of variation between groups and a p-value <0.05 was considered as statistically significant at 95% confidence interval.

Supplemental Table 1: Epidemiological details of the study subjects used in this study.

| Study groups | Total | ATB | NTB | Healthy |
|---|---|---|---|---|
| Subject (number) | 72 | 29 | 20 | 23 |
| Clinical sites (AGMC/AMC/NHAK) | 46/14/12 | 15/8/6 | 14/3/3 | 17/3/3 |
| Mean age (range) in years | 42.94 (16-83) | 41.62 (17-83) | 46.30 (25-66) | 41.43 (16-75) |
| Male (%) | 100% | 100% | 100% | 100% |
| AFB or GeneXpert (+ve/-ve/na) | 29/20/23 | 29/-/- | -/20/- | -/-/23 |
| Alcoholic (yes/no/Ex/na) | 12/38/1/21 | 1/16/-/12 | 4/12/1/3 | 7/10/-/6 |
| Smoker (yes/no/Ex/na) | 20/28/3/21 | 4/11/2/12 | 7/9/1/3 | 9/8/-/6 |
| Expectoration (yes/no/na) | 41/14/17 | 23/5/1 | 15/2/3 | 3/7/13 |
| Cough (yes/no/na) | 49/6/17 | 25/3/1 | 17/-/3 | 7/3/13 |
| Fever (yes/no/na) | 19/33/18 | 13/15/2 | 6/8/3 | -/10/13 |
| Haemoptysis (yes/no/na) | 10/46/16 | 7/22/- | 3/14/3 | -/10/13 |
| Chest Pain (yes/no/na) | 24/31/17 | 15/13/1 | 6/11/3 | 3/7/13 |
| Breathlessness (yes/no/na) | 21/33/17 | 13/14/1 | 6/11/3 | 2/8/13 |
| Wheeze (yes/no/na) | 15/38/19 | 9/18/2 | 4/12/4 | 2/8/13 |
| Haemoglobin (n) | 11.7 (29) | 11.7 (7) | 11.9 (6) | 11.59 (17) |
| Urea (n) | 26.70 (36) | 23.49 (15) | 32.1 (8) | 27.07 (13) |
| Creatinine (n) | 0.81 (35) | 0.8 (15) | 0.84 (8) | 0.81 (12) |
| Urea/Creatinine (n) | 32.43 (35) | 29.37 (15) | 38.33 (8) | 32.14 (12) |

AGMC: Agartala Government Medical College Agartala; AMC: Assam Medical College-Dibrugarh; NHAK: Nagaland Hospital Authority-Kohima; +ve: positive; -ve: negative; na: Not available; n: sample size (available data)

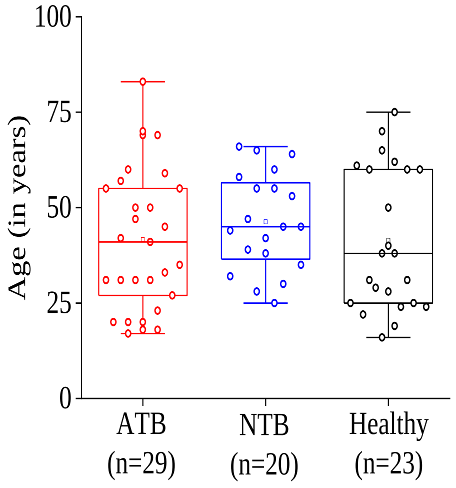


Supplementary Figure S1: Age distribution in the complete study population (n=72) including case (active tuberculosis patients: ATB) and controls (non-tuberculosis patients: NTB and healthy subjects) showed similar range.

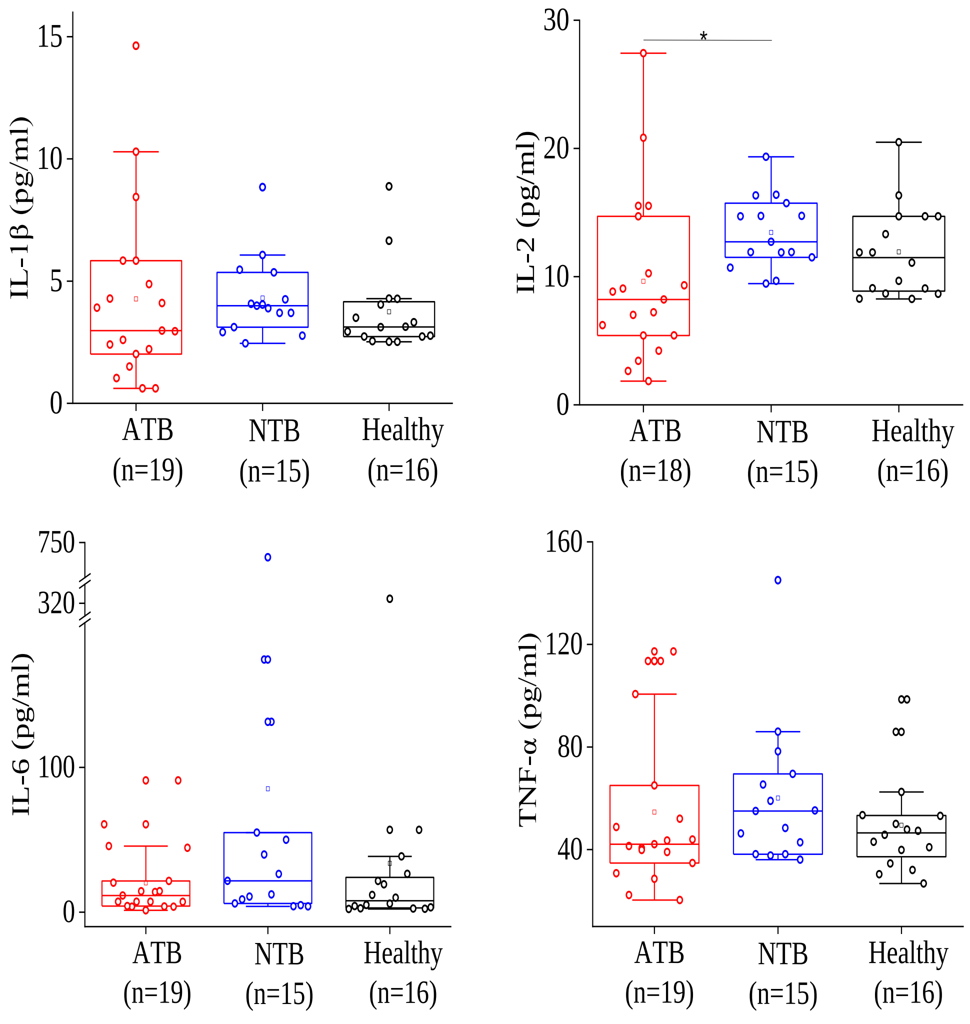


Supplementary Figure S2: Serum cytokine levels as estimated by Bioplex in active tuberculosis patients (ATB) and controls (non-tuberculosis: NTB and healthy subjects) . *: $p<0.05$

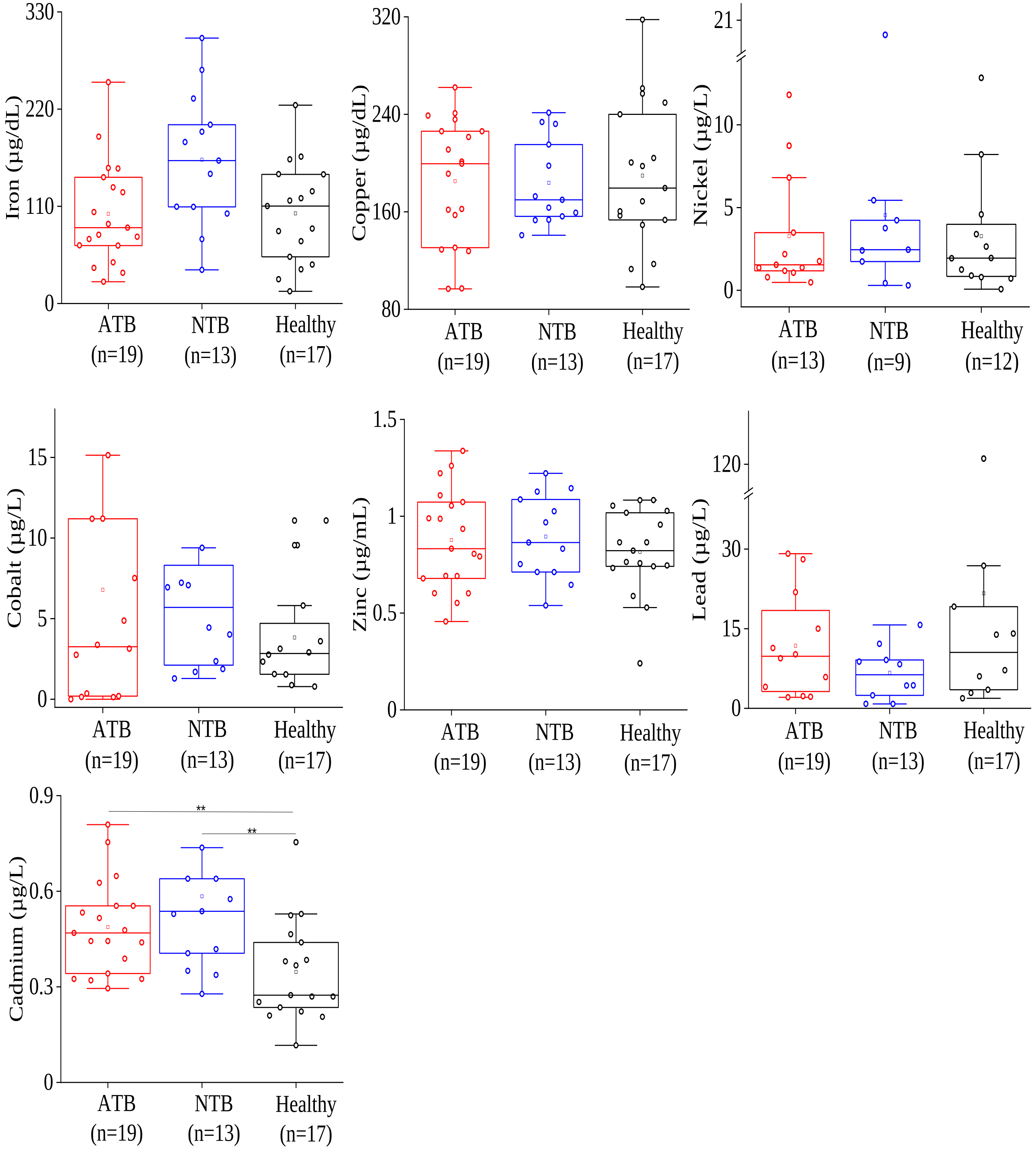


Supplementary Figure S3: Serum trace metal concentrations as estimated by Inductively coupled plasma mass spectrometry (ICP-MS) in active tuberculosis patients (ATB) and controls (non-tuberculosis: NTB and healthy subjects). *:p<0.05 ; **: p<0.01.

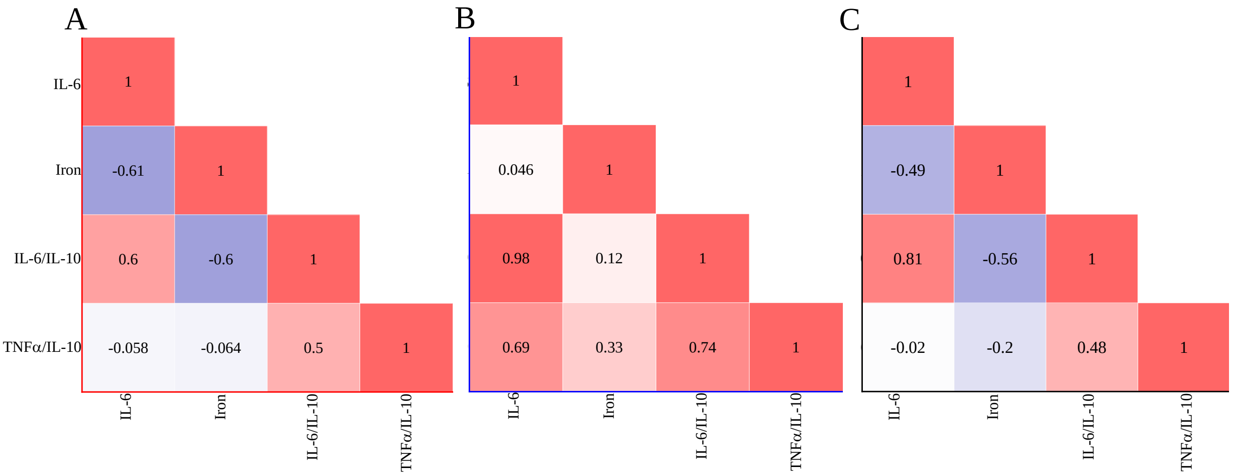


Supplementary Figure S4: Correlation (Pearson r) values between the serum IL-6 and Iron (Fe) levels in (A) active tuberculosis (ATB), (B) non tuberculosis (NTB) and (C) healthy control groups.

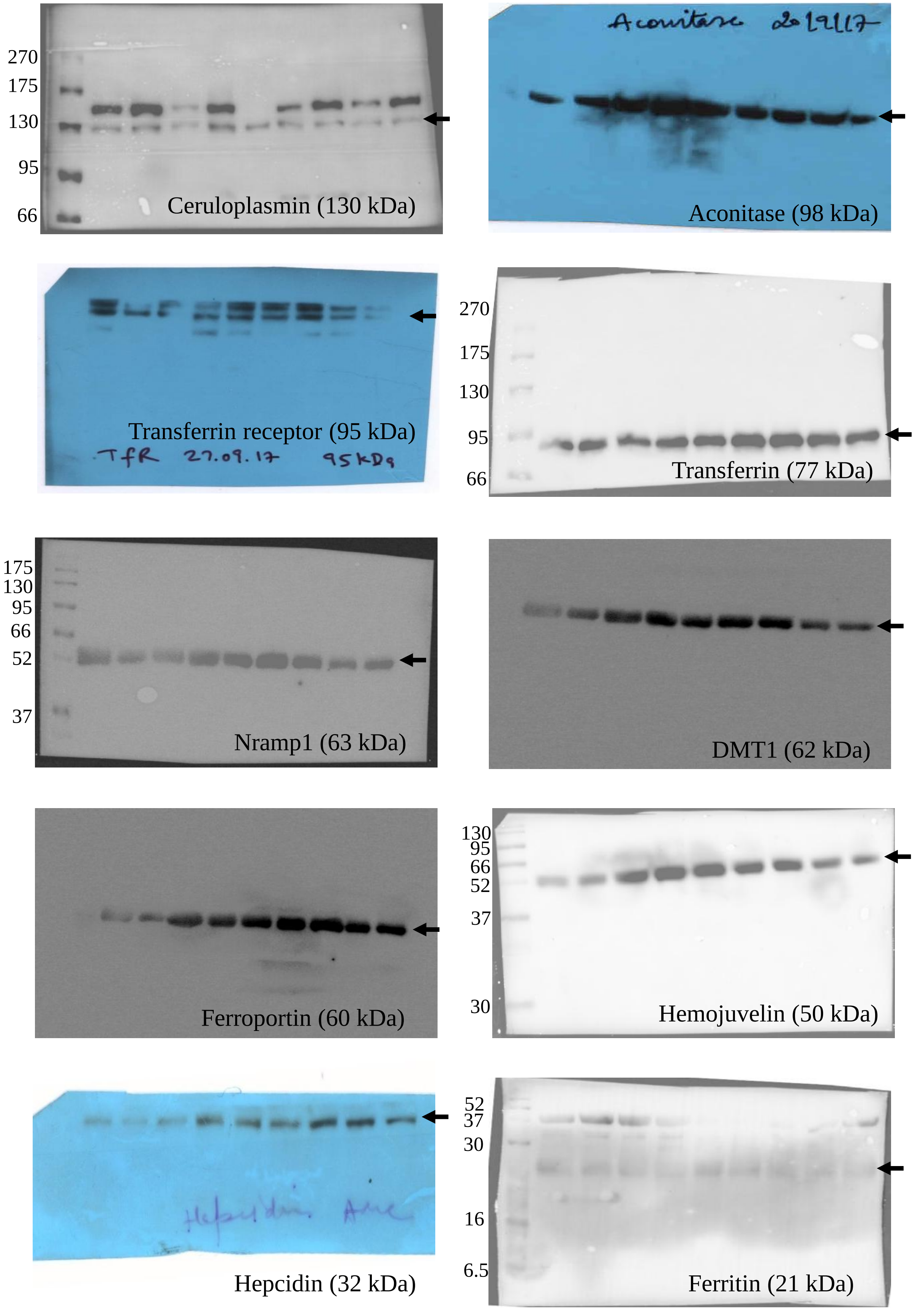


Supplementary Figure S5: Complete Western blot image of serum iron metabolizing proteins, used for intensity calculation in samples from active tuberculosis patients (ATB) and controls (non-tuberculosis: NTB and healthy subjects) and presented in Figure 2A and 2B. A representative parallel silver stained gel image is shared in Supplementary Figure S9A.

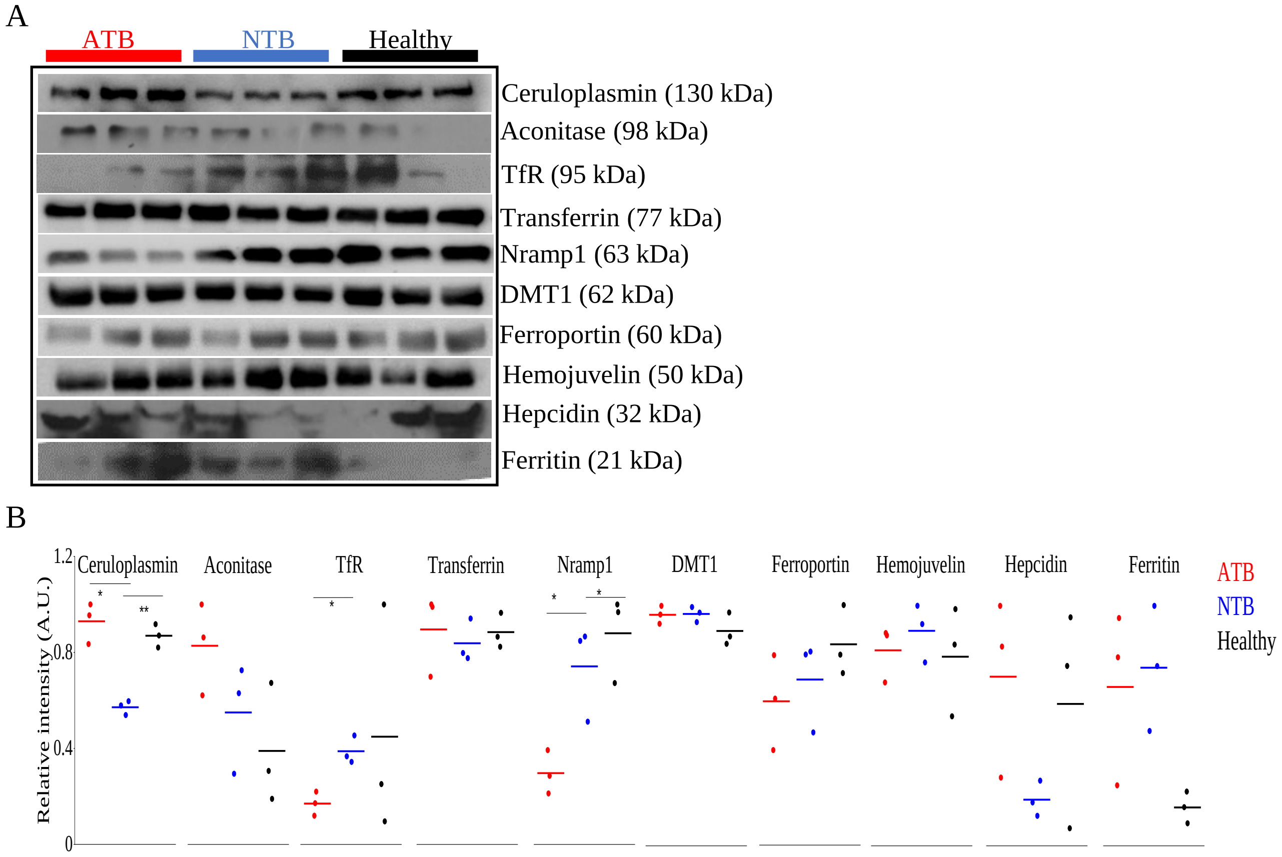


Supplementary Figure S6: Western blot images of serum iron metabolizing proteins in active tuberculosis patients (ATB) and controls (non-tuberculosis: NTB and healthy subjects) from 2nd clinical site (S6A). Complete blot images are presented in Supplementary Figure S7. Variation in intensities of individual serum protein levels in ATB, NTB and healthy control groups (S6B). TfR: Transferrin receptor; DMT1: Divalent metal ion transporter 1; *:p<0.05 ; **: p<0.01.

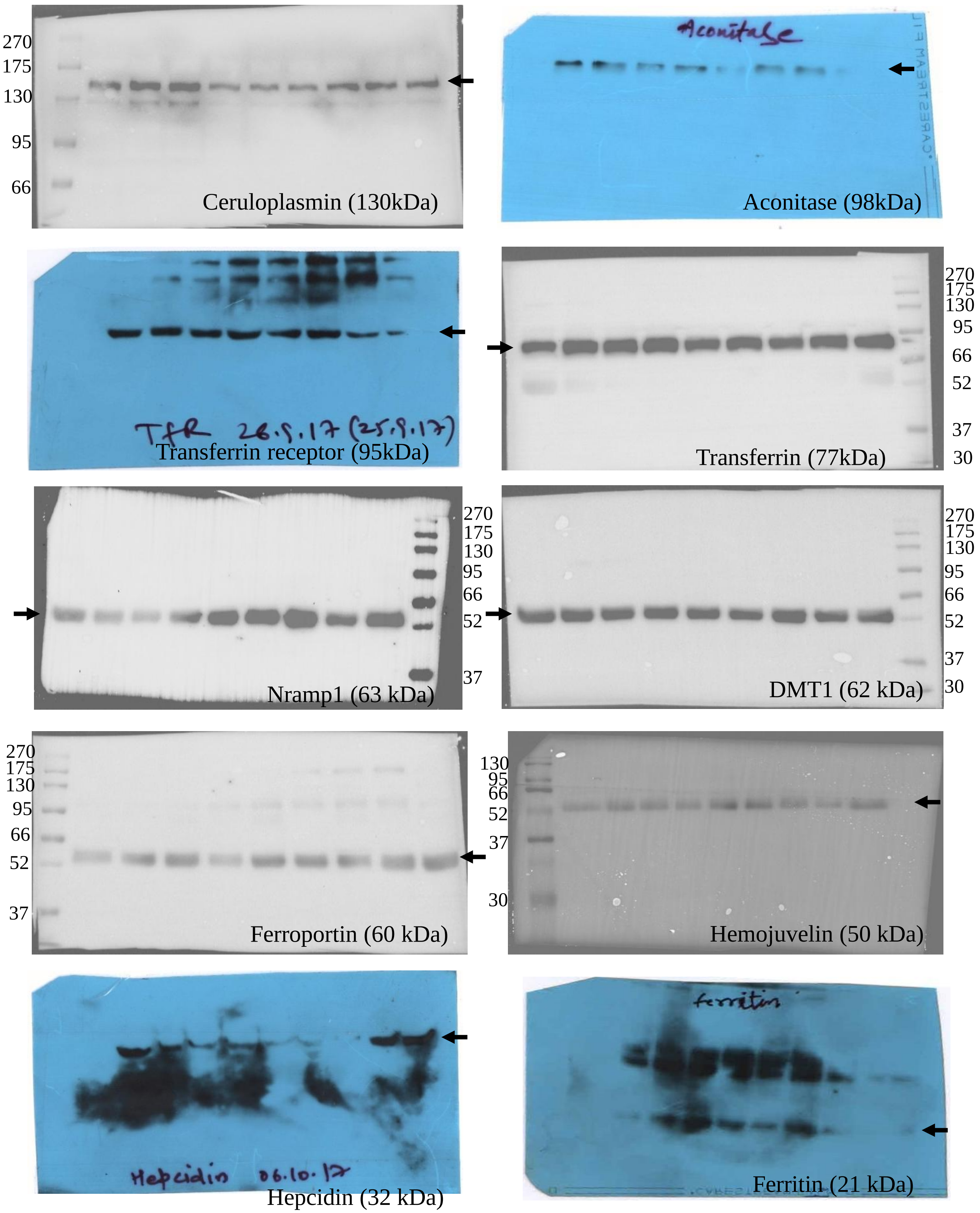


Supplementary Figure S7: Complete Western blot image of serum iron metabolizing proteins, used for intensity calculation in active tuberculosis patients (ATB) and controls (non-tuberculosis: NTB and healthy subjects) and presented in Supplementary Figure S6. A representative parallel silver stained gel image is shared in Supplementary Figure S9B.

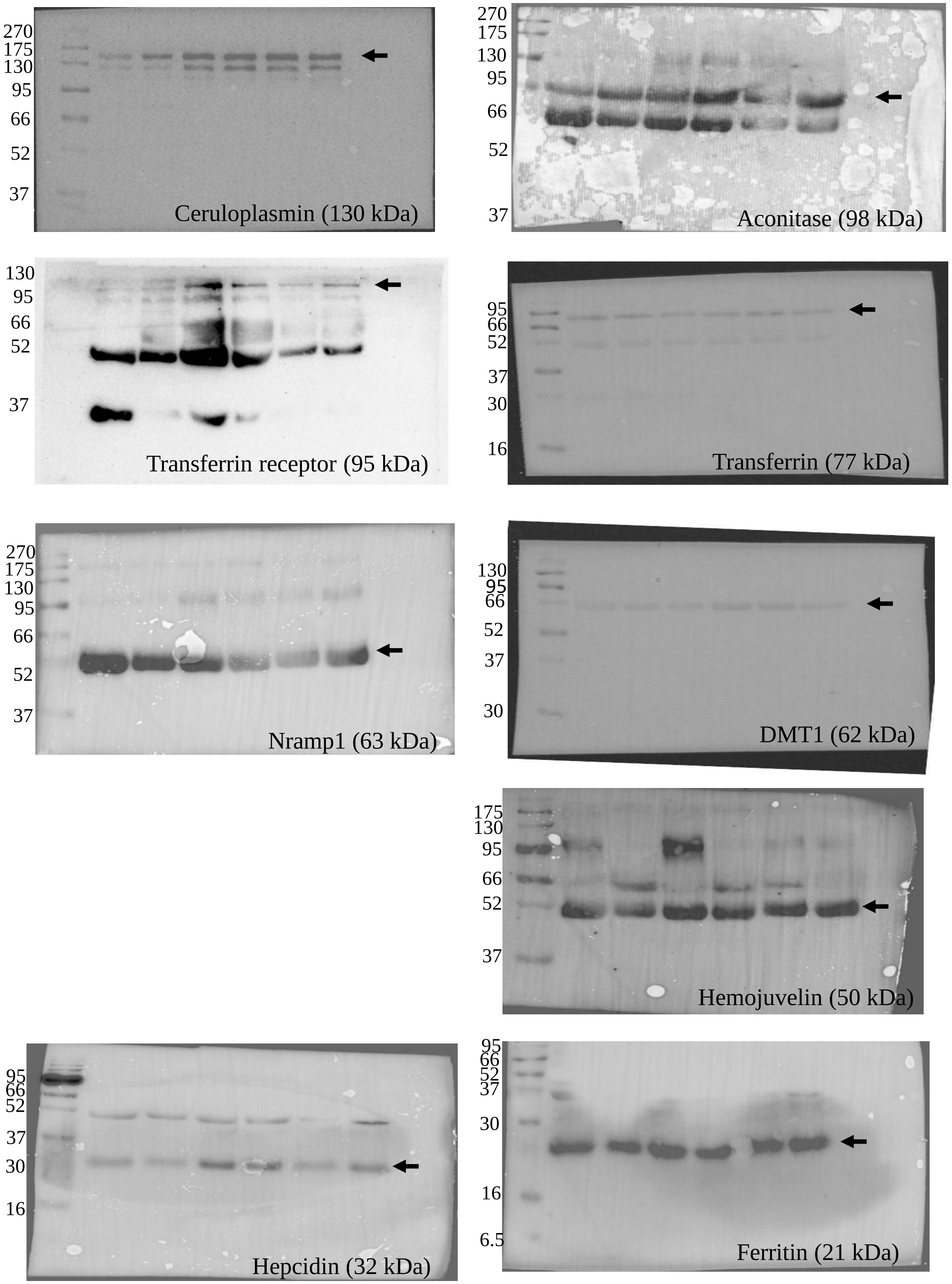


Supplementary Figure S8: Complete Western blot image of serum iron metabolizing proteins, used for intensity calculation in longitudinally followed up active tuberculosis patients (ATB) at time of presentation (0 month) and completion of treatment (6 months, clinically cured) and presented in Figure 2C and 2D. Parallel silver stained gels are presented in Supplementary Figure S10 and a representative parallel silver stained gel image is shared in Supplementary Figure S9A.

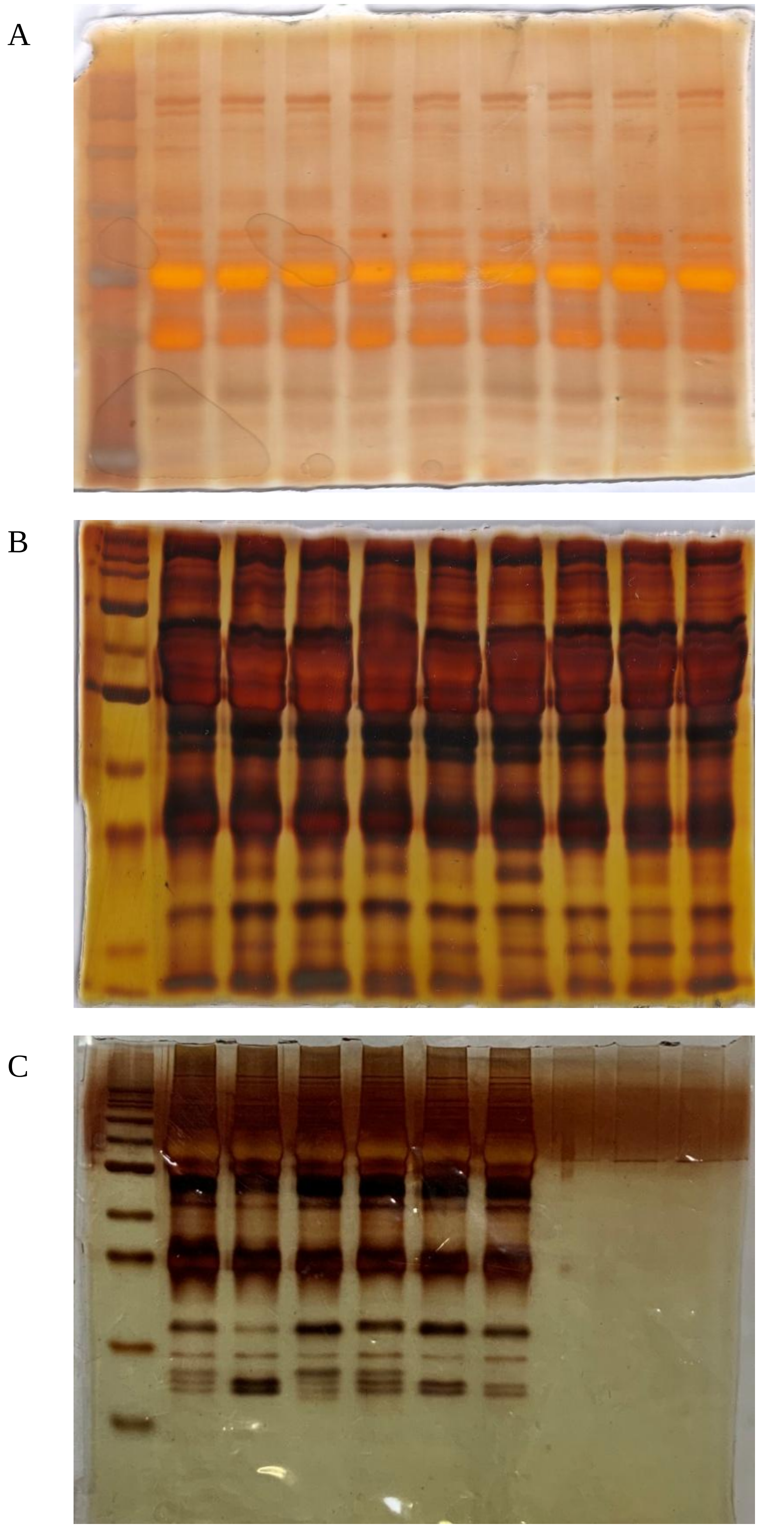


Supplementary Figure S9: Silver stained gel-images for loading control. A) Representative gel image for Supplementary Figure S5. B) Representative gel image for Supplementary Figure S7. C) Representative gel image for Supplementary Figure S8.

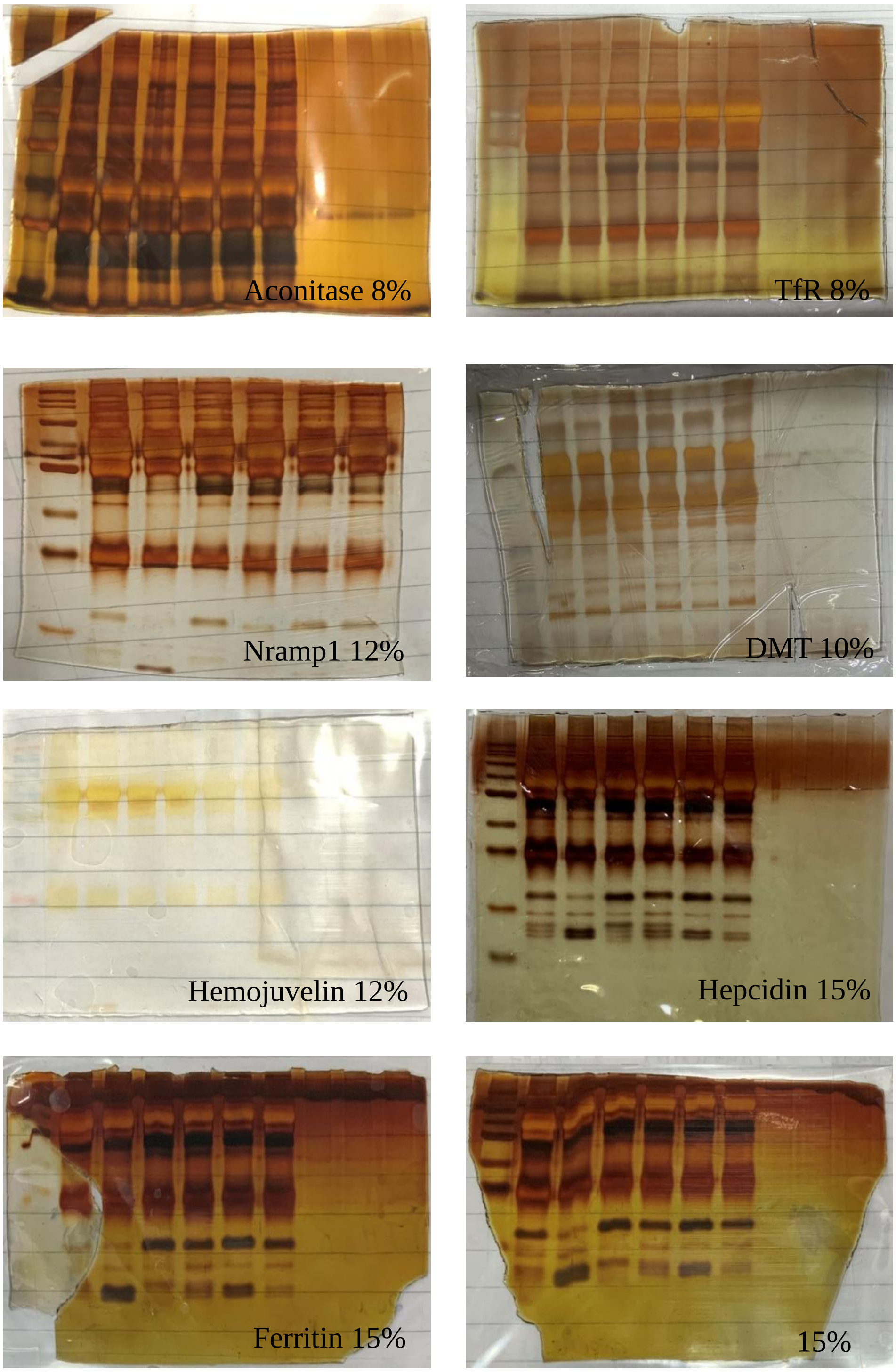


Supplementary Figure S10: Silver stained gel images of serum iron metabolizing proteins, in longitudinally followed up active tuberculosis patients (ATB) at time of presentation (0 month) and completion of treatment (6 months, clinically cured) and presented in Figure 2C, 2D, and Supplementary Figure S8.
